# Prediction of 30-Day Mortality for ICU Patients with Sepsis-3

**DOI:** 10.1101/2024.05.27.24308004

**Authors:** Z Yu, N Ashrafi, H Li, K Alaei, M Pishgar

## Abstract

**Background:** There is a growing demand for advanced methods to improve the understanding and prediction of illnesses. This study focuses on Sepsis, a critical response to infection, aiming to enhance early detection and mortality prediction for Sepsis-3 patients to improve hospital resource allocation.

**Methods:** In this study, we developed a Machine Learning (ML) framework to predict the 30-day mortality rate of ICU patients with Sepsis-3 using the MIMIC-III database. Advanced big data extraction tools like Snowflake were used to identify eligible patients. Decision tree models and Entropy Analyses helped refine feature selection, resulting in 30 relevant features curated with clinical experts. We employed the Light Gradient Boosting Machine (LightGBM) model for its efficiency and predictive power.

**Results:** The study comprised a cohort of 9118 Sepsis-3 patients. Our preprocessing techniques significantly improved both the AUC and accuracy metrics. The LightGBM model achieved an impressive AUC of 0.983 (95% CI: [0.980-0.990]), an accuracy of 0.966, and an F1-score of 0.910. Notably, LightGBM showed a substantial 6% improvement over our best baseline model and a 14% enhancement over the best existing literature. These advancements are attributed to (I) the inclusion of the novel and pivotal feature Hospital Length of Stay (HOSP_LOS), absent in previous studies, and (II) LightGBM’s gradient boosting architecture, enabling robust predictions with high-dimensional data while maintaining computational efficiency, as demonstrated by its learning curve.

**Conclusions:** Our preprocessing methodology reduced the number of relevant features and identified a crucial feature overlooked in previous studies. The proposed model demonstrated high predictive power and generalization capability, highlighting the potential of ML in ICU settings. This model can streamline ICU resource allocation and provide tailored interventions for Sepsis-3 patients.

## BACKGROUND

Sepsis [1], a life-threatening condition triggered by infection, often leads to organ failure and exhibits rapid, unpredictable progression [2]. In the United States alone, Sepsis affects approximately 1.7 million adults annually, resulting in around 270,000 deaths. Notably, recent research involving over 110,000 hospital admissions underscored a significant association between prolonged hospital stays and diminished survival rates, particularly for stays exceeding nine days. Globally, Sepsis accounted for nearly a fifth of all reported fatalities in 2017, with an estimated 11 million deaths out of nearly 49 million reported cases [3]. Given the severity of Sepsis’s impact, understanding the factors contributing to elevated mortality rates among patients is imperative.

The understanding of Sepsis has evolved notably with the introduction of Sepsis-3 by the Third International Consensus Definitions for Sepsis and Septic Shock in 2016 [4]. This new paradigm, emphasizing a clearer correlation between infection and subsequent organ failure, calls for fresh avenues of research. It not only reshapes diagnostic and treatment approaches but also provides clinicians and researchers with a refined framework for identifying and analyzing Sepsis cases accurately. Familiarity with this contemporary approach is indispensable for the development of effective diagnostic and therapeutic strategies, empowering healthcare professionals to confront this formidable medical challenge more effectively.

Previously, methods for assessing Sepsis severity and mortality risk relied heavily on tools like the Simplified Acute Physiology Score-II (SAPS-II) [5], a severity-of-disease classification system primarily based on physiological data collected within the first 24 hours of ICU admission. However, the limitations of SAPS-II, particularly its susceptibility to missing data and rapid changes in patient condition post-admission, pose challenges in the dynamic ICU setting. Other methods, such as calculating a Sequential Organ Failure Assessment (SOFA) score [6], may suffer from subjectivity and interrater variability, leading to inconsistent results.

In addition to traditional scoring methods, conventional statistical models like Logistic Regression [7] have been widely used for outcome prediction in Sepsis. However, these models often struggle to capture the intricate, non-linear relationships inherent in medical data. Moreover, they rely on assumptions about data distribution that are rarely met in medical contexts, leading to suboptimal predictions. The inadequacy of these models underscores the need for more advanced analytical techniques.

Machine learning (ML) models have shown great promise in capturing complex patterns. [8–10]. In recent years, ML models have emerged as effective alternatives, particularly for handling high-dimensional and unnormalized data [11–16]. Due to its unique characteristics, such as efficiency, accuracy, and the ability to handle large datasets, LightGBM [17] stands out. Leveraging those ensemble learning techniques, LightGBM sequentially builds decision trees to correct errors and improve predictive performance. Despite the increasing use of ML algorithms to predict mortality in ICU patients with Sepsis [18,19], none have yielded satisfactory results, potentially due to poor feature selection methodologies and inadequate parameter tuning.

To address these challenges, our proposed model incorporates a novel decision tree-based Entropy Analysis [20] for feature selection, identifying significant factors for mortality prediction. This approach enhances computational efficiency and identifies hidden relationships in complex datasets, offering a more nuanced and precise approach to medical prediction.

Our study aims to demonstrate the clinical applicability of this feature engineering process and evaluate the predictive performance of proposed model in mortality prediction for Sepsis-3 patients. Additionally, the learning curve of our proposed model is plotted to validate its generalization and predictive accuracy. Our prediction model complies with the standards of the Transparent Reporting of a Multivariable Prediction Model for Individual Prognosis or Diagnosis (TRIPOD) initiative, guaranteeing thorough and transparent reporting [21,22]. The graphical abstract of this research is described in Figure 1.

**Figure 1:**
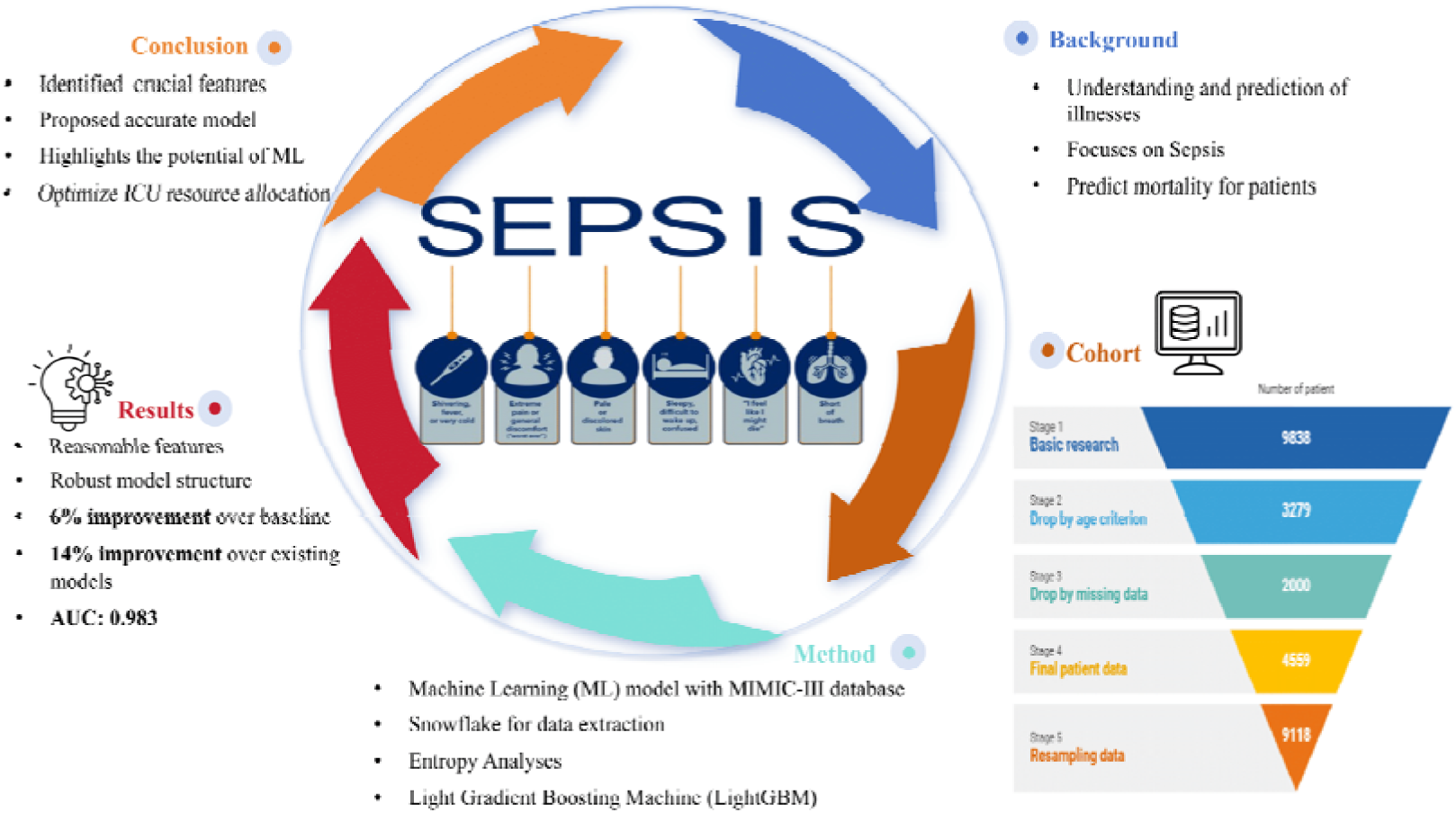
A graphical abstract of this study.

## METHODS

### Data Availability

The Medical Information Mart for Intensive Care III (MIMIC-III) is a comprehensive dataset, available to the public via https://physionet.org/content/mimiciii/1.4/, which includes de-identified health information from more than 40,000 ICU admissions at the Beth Israel Deaconess Medical Center from 2001 to 2012 [23]. Created by the MIT Lab for Computational Physiology, MIMIC-III encompasses diverse data categories such as demographics, vital signs, laboratory test results, medications, and mortality outcomes. This extensive dataset enables multifaceted research in clinical informatics.

### Patient Selection

We initially included patients who were classified as “Sepsis,” “severe Sepsis,” and “septic shock.” To exclude incomplete and repeated data, we have further narrowed data, as illustrate in Figure 1, adhering to specific inclusion criteria: (I) patients aged 18 years or older; (II) patients with complete demographic and lab test results and with fewer than 20% of features missing; (III) patients with SOFA scores.

**Figure 1:**
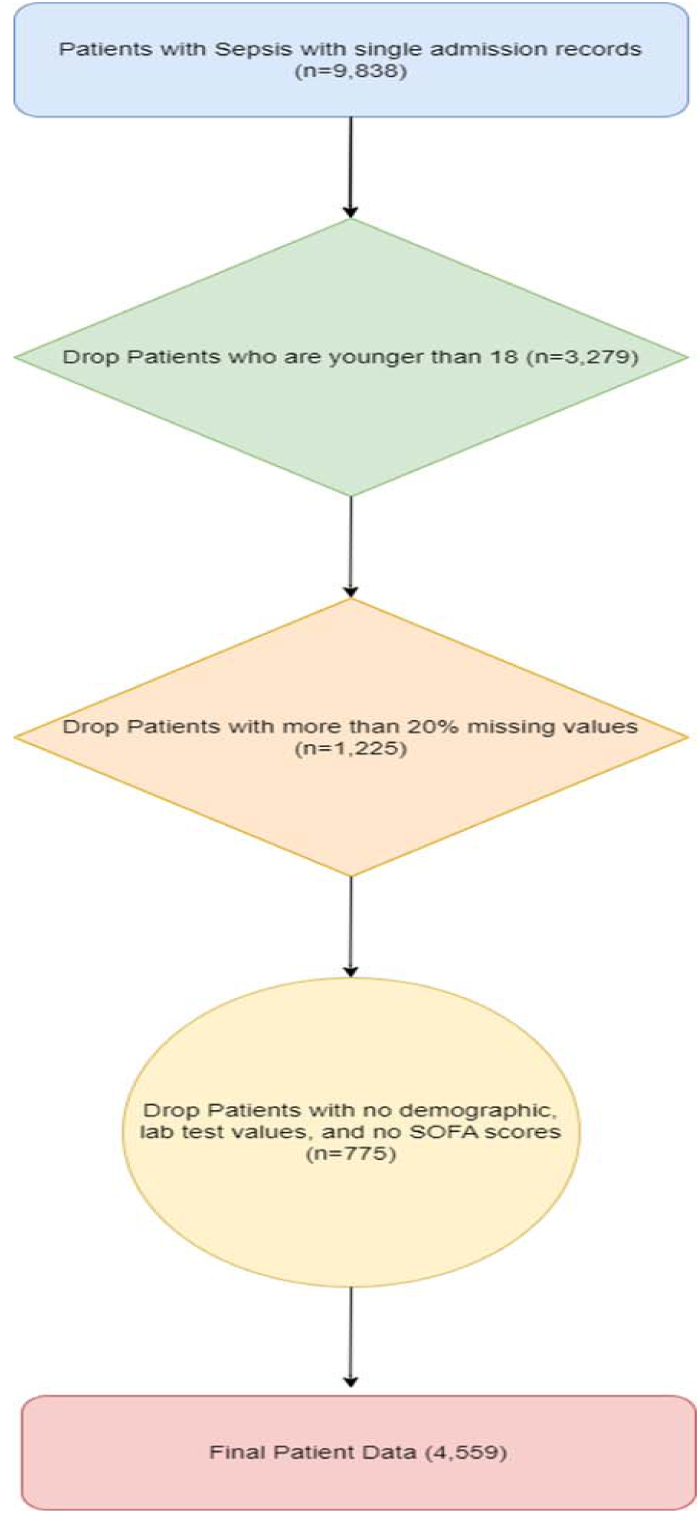
It illustrates the process of patient selection.

Patients with more than 20% of features missing were excluded by calculating the percentage of missing values for each patient and removing those who exceeded this threshold. For the remaining patients, missing values were addressed using random sampling imputation, where missing values were replaced with random values drawn from the existing non-missing values in the same column. This technique helps preserve the data distribution and maintain variability within the dataset, mitigating biases and minimizing information loss inherent in incomplete datasets [24]. Moreover, this approach is particularly effective when the proportion of missing data is relatively low, as it helps ensure the imputed values are representative of the observed values. Sensitivity analyses were conducted to evaluate the impact of this imputation method, confirming that it did not introduce significant bias and maintained the robustness and predictive power of our models. Additionally, this method generates additional samples for model training, enhancing the model’s robustness and predictive power. Duplicate records were identified and removed based on unique patient identifiers. For patients with multiple admissions, only the first admission record was retained to avoid redundancy and potential bias.

### Feature Selection and Pre-processing

The feature selection process of the research unfolds in two distinct stages. Firstly, Entropy Analysis was applied using importance scores derived from decision trees, which measure each feature’s contribution to reducing uncertainty (entropy) in the prediction model. Features with higher information gain scores were prioritized as they indicated greater predictive power. To set a threshold, we conducted an initial grid search to determine the approximate ranges of information gain values. We decided to include features that provided at least 13% of the maximum importance score (approximately 0.01), which resulted in retaining the top 30 features. This threshold was selected to balance predictive power and model complexity. The effectiveness of this threshold was validated through experimental results and cross-validation, ensuring that the retained features offered the most predictive power while maintaining manageable model complexity. Secondly, clinical experts specializing in critical care reviewed and refined our selected features, ensuring their practical relevance and significance in the ICU setting. Their input was instrumental in adding certain features based on their medical knowledge and experience, while also recommending the removal of less significant features.

Initially, drawing from existing literature and expert insights, demographic data including age, gender, ethnicity, weight, height, and body mass index (BMI), along with hospital and ICU lengths of stay and in-hospital mortality status, were extracted from initial ICU admission records. Vital signs such as heart rate (HR), mean arterial pressure (MAP), temperature (TEMP), respiratory rate (RR), and oxyhemoglobin saturation (SpO2) were recorded from the first 24 hours of ICU admission. Additionally, laboratory values encompassing blood routine examination, liver and kidney function, blood glucose, and arterial blood gas (ABG) measurements were abstracted. Given the high sampling frequency, maximum, minimum, and mean values were utilized to incorporate vital signs and related laboratory indicators effectively.

Subsequently, employing Entropy Analysis based on Decision Trees with a threshold of 30, we refined the feature set, resulting in 30 features selected for further analysis as shown in Table 1. Furthermore, owing to the constraints posed by the relatively modest final sample size (4,559), we implemented bootstrapping [25], a statistically robust resampling technique aimed at augmenting the volume and diversity of the original patient population. Bootstrapping creates a larger dataset by repeatedly sampling with replacement, enhancing statistical power and model robustness. However, this technique has limitations, such as the risk of overfitting, where models perform well on training data but poorly on unseen data, and increased model variance, which affects prediction stability. To mitigate these issues, we recommend combining bootstrapping with other data augmentation techniques and cross-validation methods, and validating models on external datasets to ensure robustness and generalizability.

**Table 1:**
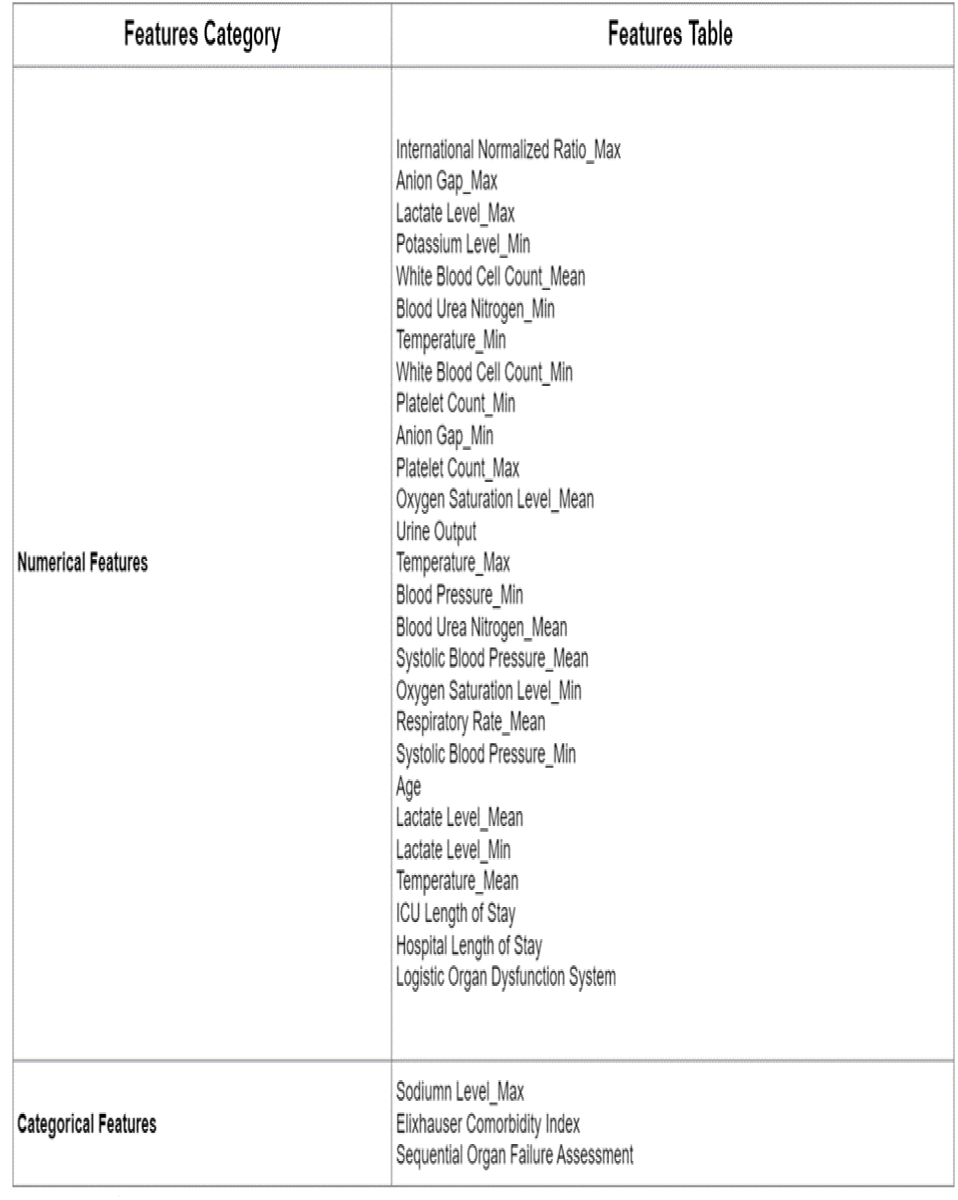
A summary of numerical and categorical features.

| Features Category | Features Table |
| --- | --- |
| Numerical Features | International Normalized Ratio_Max<br>Anion Gap_Max<br>Lactate Level_Max<br>Potassium Level_Min<br>White Blood Cell Count_Mean<br>Blood Urea Nitrogen_Min<br>Temperature_Min<br>White Blood Cell Count_Min<br>Platelet Count_Min<br>Anion Gap_Min<br>Platelet Count_Max<br>Oxygen Saturation Level_Mean<br>Urine Output<br>Temperature_Max<br>Blood Pressure_Min<br>Blood Urea Nitrogen_Mean<br>Systolic Blood Pressure_Mean<br>Oxygen Saturation Level_Min<br>Respiratory Rate_Mean<br>Systolic Blood Pressure_Min<br>Age<br>Lactate Level_Mean<br>Lactate Level_Min<br>Temperature_Mean<br>ICU Length of Stay<br>Hospital Length of Stay<br>Logistic Organ Dysfunction System |
| Categorical Features | Sodium Level_Max<br>Elixhauser Comorbidity Index<br>Sequential Organ Failure Assessment |

The Min-Max Scaler was employed to rescale numerical features, thereby normalizing them to a range of 0 to 1. This technique was chosen because it preserves the relationships between the original data points and is effective when the data does not follow a Gaussian distribution. This procedure plays a crucial role in ensuring that all numerical features contribute equally to the analysis, thereby mitigating biases resulting from features with larger scales. Categorical features underwent transformation using Label Encoder, which involves converting categorical labels into numerical codes, thereby enabling their integration into regression and ML models. This method was selected for its simplicity and efficiency, particularly suitable for tree-based models and manageable numbers of unique categories. These preprocessing techniques serve to standardize the dataset, a fundamental prerequisite for efficient model training and evaluation. Furthermore, they ensure that analyses accurately reflect the original measurements and categories present in the dataset. In conclusion, the complete process is demonstrated in Figure 2.

**Figure 2:**
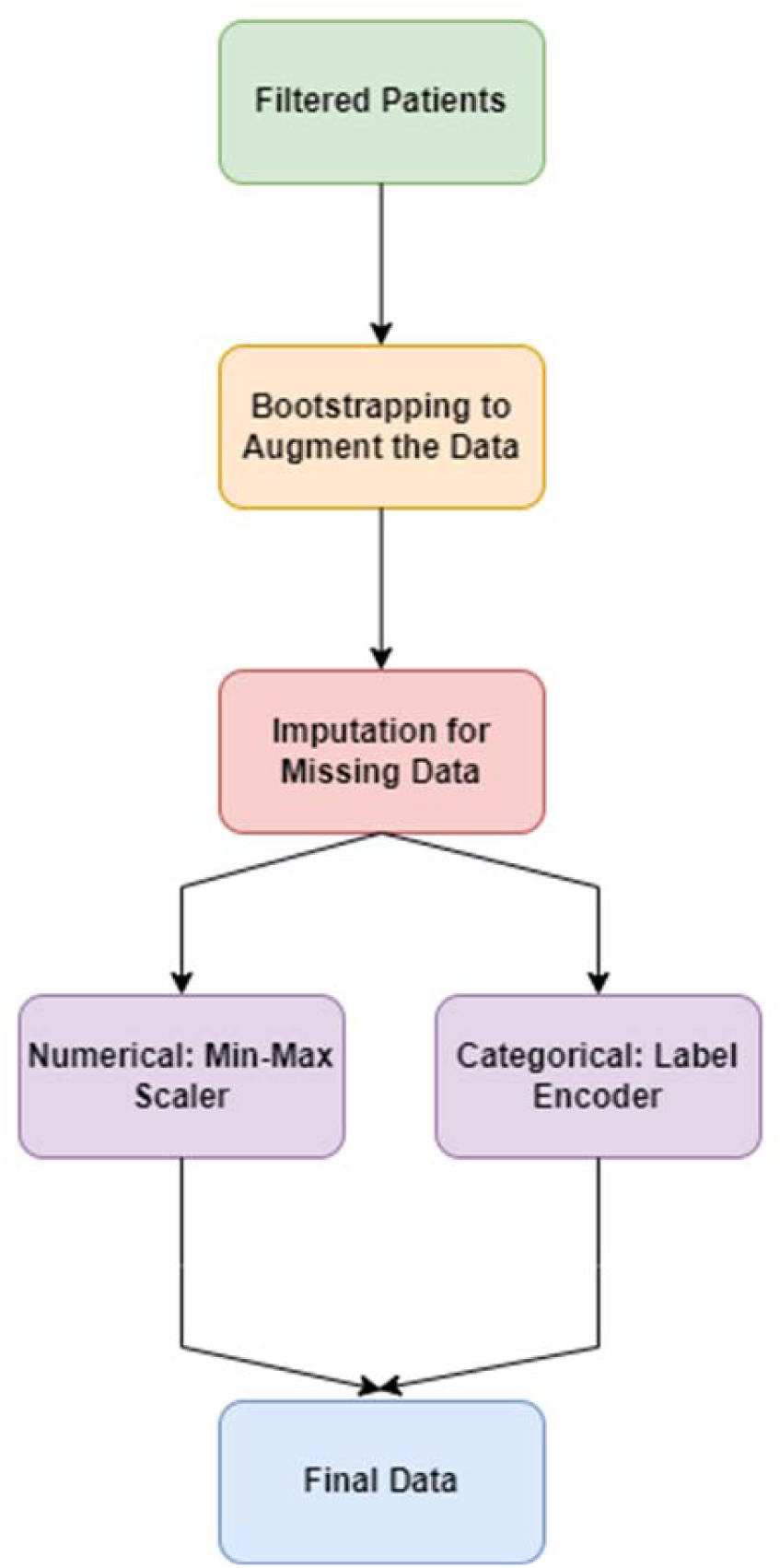
It presents the work flow of data preprocessing.

### Model Development and Optimization

Our final dataset encompasses 9,118 patients with 30 features. A train-test split is executed with an 80/20 ratio to facilitate model evaluation. To mitigate overfitting, we utilize Grid Search CV to identify the optimal combination of hyperparameters. In the GridSearchCV, the LGBMClassifier was tuned using several key parameters: num_leaves, which controls the complexity of the tree and thus influences the model’s ability to capture features and avoid overfitting; learning_rate, which determines the step size at each iteration while moving towards a minimum of the loss function; feature_fraction, representing the fraction of features used for training each tree; bagging_fraction, indicating the fraction of data used for each iteration of bagging; and bagging_freq, defining how frequently bagging is performed. The cross-validation (cv) value was set to 5, meaning that the data was split into five subsets, with each subset used as a validation set once while the others were used for training. This ensures that the model is evaluated on different data portions, improving its generalization ability. Due to time constraints, we initially conducted a coarse GridSearch to determine the approximate ranges for these parameters. The final parameter ranges were set as follows: num_leaves [31, 50, 100], learning_rate [0.05, 0.1, 0.3], feature_fraction [0.8, 0.9, 1.0], bagging_fraction [0.7, 0.8, 0.9], and bagging_freq [3, 5, 7]. After running GridSearchCV, the optimal parameters identified were num_leaves at 100, learning_rate at 0.3, feature_fraction at 0.8, bagging_fraction at 0.8, and bagging_freq at 3. These parameters collectively enhance the model’s performance by balancing complexity, learning pace, and robustness through feature and data fraction adjustments.

We construct various ML algorithms, including Logistic Regression, LightGBM, CatBoost [26], Random Forest [27], K-Nearest Neighbors (KNN) [28], Support Vector Machine (SVM) [29], and Extra Gradient Boosting (XGBoost) [30]. The training process of these models included a grid search of model parameters. This search process aimed to find the best model which was determined based on the Area Under the Receiver Operating Characteristic (AUROC) scores of the cross-validation cohort. Accuracy and F1 scores are also computed for comparative analysis of model performance. Given the widespread adoption of AUC as an evaluation metric in existing literature, the selection of the proposed model is based on AUC on the cross-validation. LightGBM emerges as the top performer, consistent with our expectations of its superior performance compared to other models. LightGBM includes Gradient-based One-Side Sampling (GOSS), which improves computational efficiency by focusing on the most significant instances, and Exclusive Feature Bundling (EFB), which reduces feature dimensionality by bundling mutually exclusive features. Additionally, LightGBM is known for its speed and efficiency, with fast training times and low memory usage, making it ideal for large datasets. It also efficiently handles sparse data, which is common in real-world datasets. These features make LightGBM particularly effective for handling large-scale data and complex feature interactions. Since we observed a notably high importance score for the feature ho HOSP_LOS during Entropy Analysis, we calculated the AUC scores of the proposed model with and without this feature. An overview of the methodologies employed is illustrated in Figure 3.

**Figure 3:**
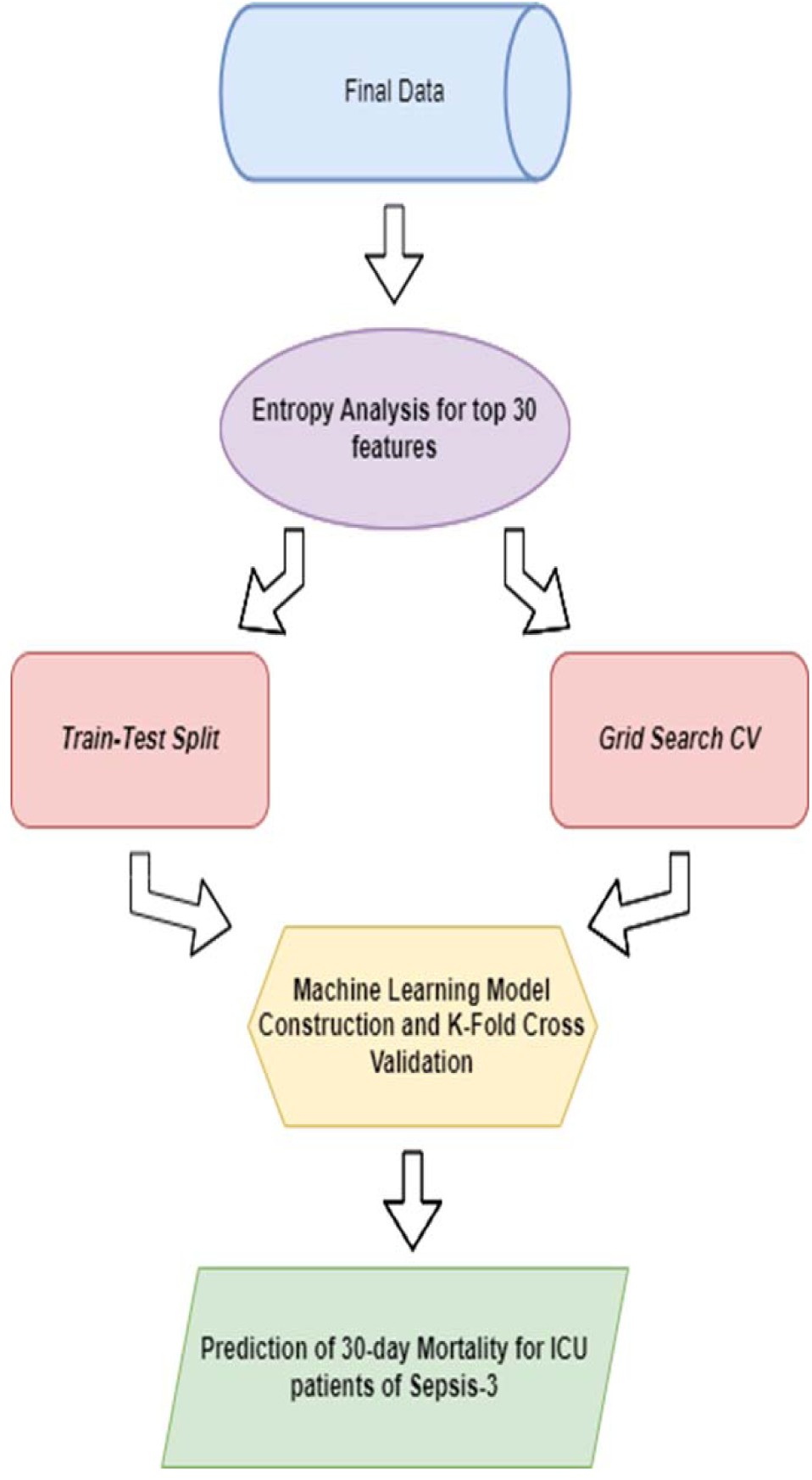
It elaborates our novel feature engineering methodologies and model construction process.

### Statistical Analysis of Models

To validate the statistical robustness of our model results, we employed comprehensive statistical tests, utilizing diverse criteria to evaluate overall performance.

To ascertain whether these AUC scores were statistically different, we conducted the Mann-Whitney U Test (Wilcoxon Rank-Sum Test) [31]. Unlike the Student’s t-test, the Mann-Whitney U Test does not require assumptions about the underlying dataset distribution, making it more suitable for our analysis. This non-parametric test is preferred over other non-parametric tests like the Kruskal-Wallis test because it is specifically designed to compare two independent samples, making it ideal for comparing AUC scores. The null hypothesis posits that the AUC scores with and without HOSP_LOS are not statistically different, while the alternative hypothesis suggests AUC scores are significantly different.

Lastly, we conducted a statistical analysis on our train/validation dataset to compare their cumulative distributions. Utilizing the Kolmogorov-Smirnov test for its non-parametric nature, we made no assumptions about the specific distribution of the data [32]. This test was chosen over parametric tests like the Chi-Square test because it compares the entire distributions of two samples and is sensitive to differences in both location and shape of the empirical cumulative distribution functions. This is particularly important as some features in our dataset may not adhere to a normal distribution. With a predetermined significance level of 0.05, our null hypothesis assumes no statistically significant difference between the test and validation sets.

### Feature Impacts

To deepen our analysis, we utilized SHapley Additive exPlanations (SHAP) [33] analysis to evaluate feature importance and elucidate the decision-making mechanisms of the predictive models, particularly within the framework of random forests. This advanced technique quantifies the influence of each feature on the model’s predictions, offering valuable insights into the reasoning behind specific predictions. SHAP values were calculated by computing the average marginal contribution of a feature value across all possible combinations of features. This provides a unified measure of feature importance, reflecting the impact each feature has on the model’s output.

## RESULTS

### Cohort Characteristics Model Completion

Following our approach for feature selection and data augmentation for ICU patients with Sepsis-3 discussed before in this paper, our final dataset contained 9118 patients from MIMIC-III database. The selected cohort was then split into train/test cohorts randomly with a ratio of 80/20, which yielded a result of 7294 patients for the train and 1824 patients for the test cohorts. In addition to mortality rates, we analyzed demographic details such as age, gender, and ethnicity, and clinical characteristics including comorbidities, vital signs, and laboratory test results. The average age of the cohort was 65.13 years, with a standard deviation of 17.67 years. The gender distribution was 56.6% male and 43.4% female. Ethnicity was diverse, with the majority being White (70.9%), followed by Black/African American (7.3%), and other ethnicities (21.8%). Clinical characteristics included comorbid conditions such as diabetes (28.0%). Vital signs recorded included mean arterial pressure (min: 54.74, max: 106.11, mean: 76.17), heart rate (min: 73.09, max: 107.55, mean: 88.44), and respiratory rate (min: 12.82, max: 28.20, mean: 19.92), while laboratory tests included measurements of white blood cell count (min: 10.89, max: 15.41, mean: 13.14), hemoglobin (min: 10.00, max: 11.94), and creatinine levels (min: 1.36, max: 1.73).

The training cohort was used to train the model, with Grid Search CV and K-fold cross-validation identifying optimal parameters and validating the model. The testing cohort evaluated model performance. The best model was chosen based on AUC performance on the test set. Mortality rates were 19.5% for the training cohort (1422 out of 7294) and 20% for the testing cohort (365 out of 1824). The Mann-Whitney U Test for AUC results of LightGBM with and without HOSP_LOS yielded a P-value of 9.182598395744396e-89, indicating significantly different AUC scores. The Kolmogorov-Smirnov test compared training and test data distributions, with a predetermined significance level of 0.05. The P-value for HOSP_LOS was 0.1062, indicating no significant differences between cohorts. For the Logistic Organ Dysfunction System (LODS), the P-value was 0.1648, also showing no significant differences. However, P-values for age and average white blood cell counts (WBC_mean) were below the 0.05 threshold, indicating statistically significant differences in distribution between the training and test cohorts.

The statistically significant differences in age and WBC_mean between cohorts indicate potential variations in the patient populations, affecting model generalizability. Age differences imply influence on mortality predictions, given age’s critical role in Sepsis-3 outcomes. Variations in WBC_mean highlight different inflammatory responses, impacting predictions. These findings suggest stratifying data by age and WBC_mean or using techniques like propensity score matching to balance these covariates, ensuring robust model performance across diverse populations. Details are illustrated in Table 2, which indicates no statistically differences between cohorts, except for age and WBC_mean. This ensures robust model generalization and performance across datasets.

**Table 2:**
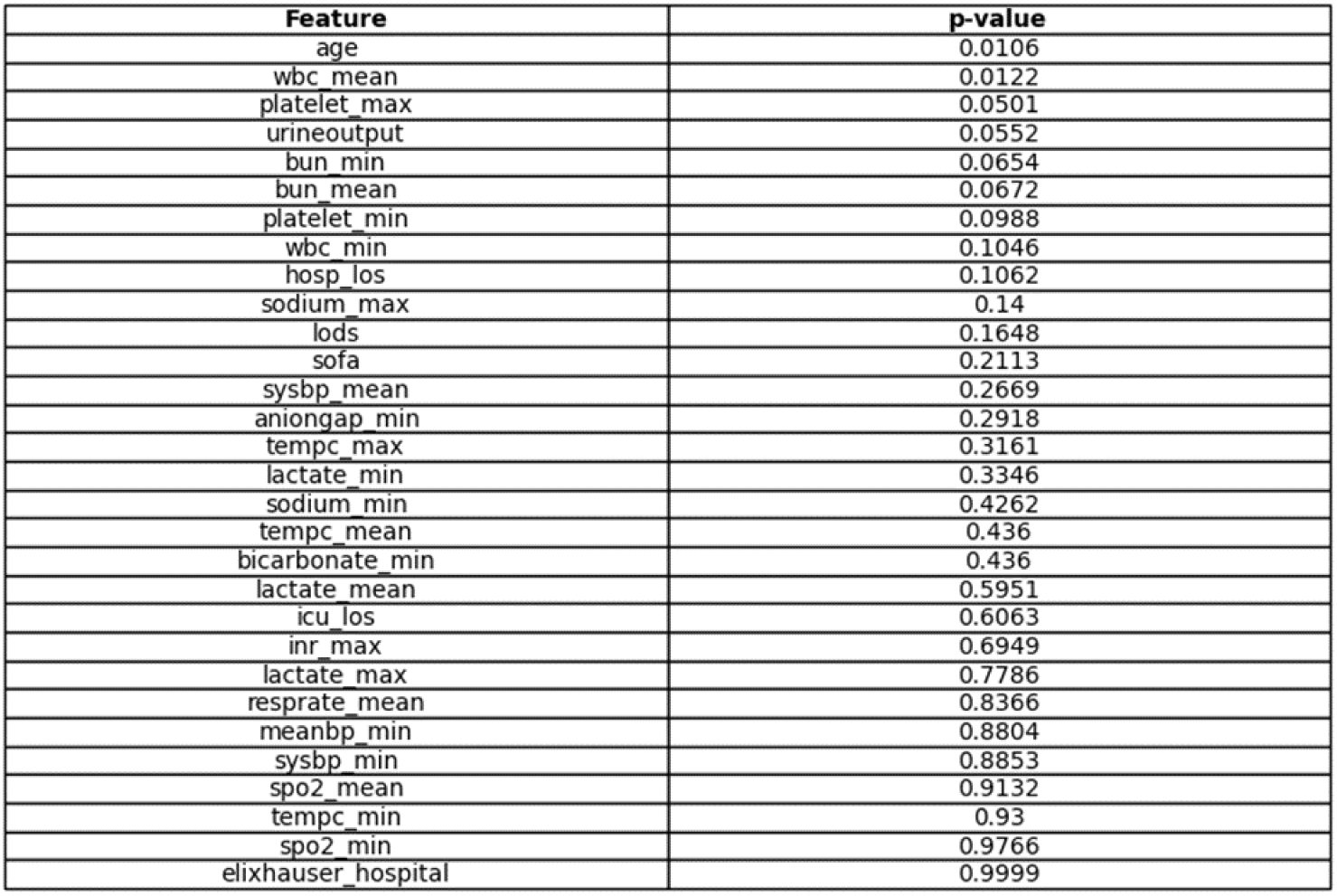
It illustrates the P-values of train/test cumulative distributions for all the significant features.

### Evaluation Metrics, Proposed and Baseline Models Performance

The summary of the results for both proposed and baseline models are shown in Table 3. The proposed approach resulted in the following metrics, AUC = 0.983, 95% CI = [0.980-0.990] as shown in Figure 4, accuracy score = 0.966, F1 score = 0.910, underscoring not only its accuracy but also its robustness in minimizing both Type I and Type II errors, thereby affirming its suitability for our predictive modeling tasks.

**Table 3:** A summary of various evaluation metrics for the models.

|  | LightGBM | Random Forest | KNN | CatBoost | XGB | SVM | Logistic Regression |
| --- | --- | --- | --- | --- | --- | --- | --- |
| AUC | 0.983 | 0.926 | 0.863 | 0.853 | 0.840 | 0.832 | 0.661 |
| Accuracy Score | 0.966 | 0.968 | 0.840 | 0.936 | 0.926 | 0.837 | 0.846 |
| F1 Score | 0.910 | 0.915 | 0.546 | 0.818 | 0.790 | 0.339 | 0.479 |

**Figure 4:**
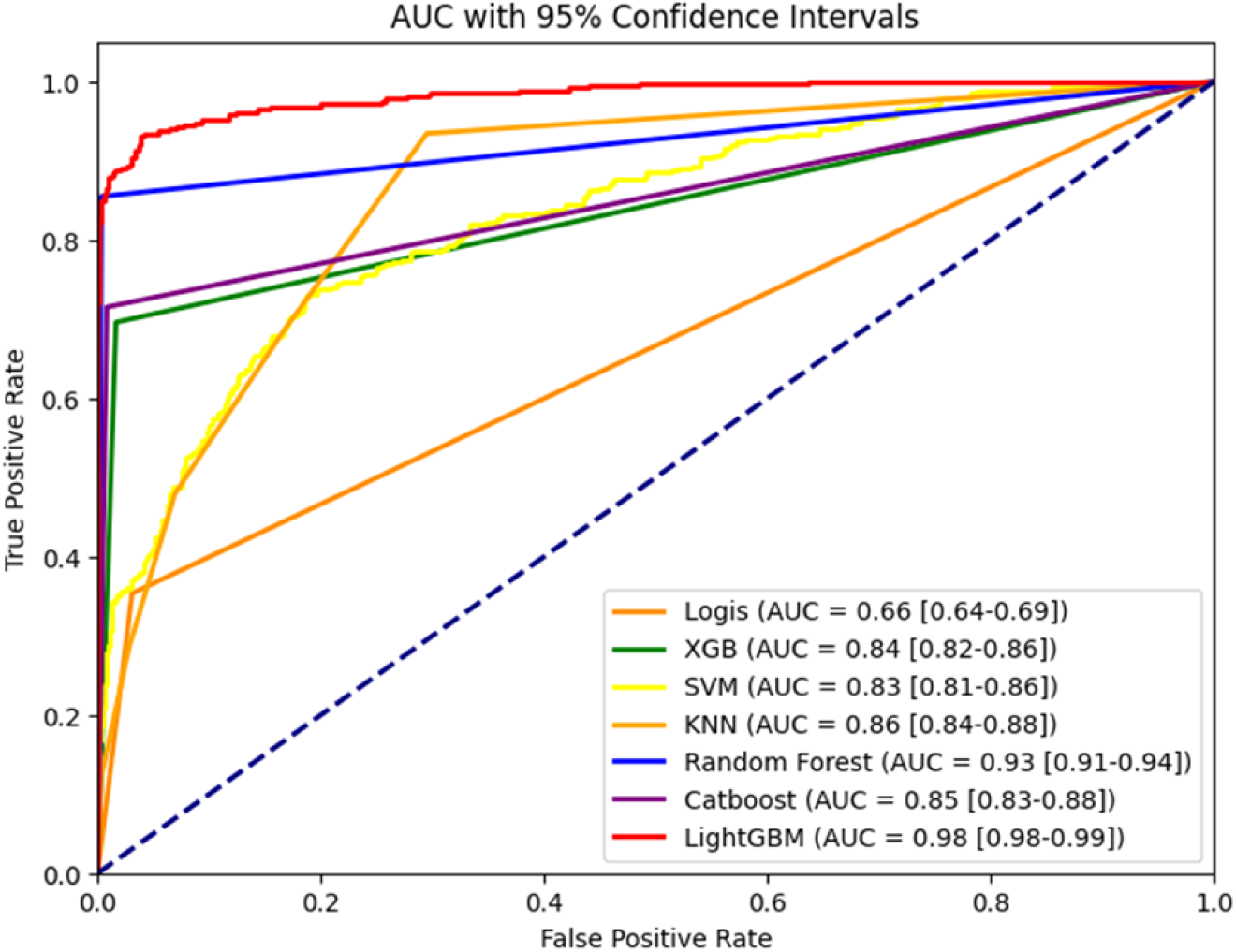
A comparison of the ROC AUCs for all Models.

In addition to the AUC score, we also evaluated the LightGBM model using precision, recall, and the confusion matrix to offer a more comprehensive evaluation of model performance. For the LightGBM model, the precision is 0.9618, indicating that 96.18% of the instances predicted as positive were indeed positive. The recall is 0.8195, meaning that 81.95% of the actual positive instances were correctly identified by the model. The F1 score, which balances precision and recall, is 0.8850, reflecting a strong overall performance. The confusion matrix further details the model’s predictions, with 1475 true negatives, 277 true positives, 11 false positives, and 61 false negatives. This indicates that the model has a low false positive rate and a relatively low false negative rate, contributing to its high precision and recall values. Finally, the R-squared value for the LightGBM model is 0.7385, suggesting that approximately 73.85% of the variance in the dependent variable is explained by the model. These additional metrics collectively demonstrate the robust performance of the LightGBM model across various evaluation criteria.

On the other hand, the baseline model development utilizing the MIMIC-III database, Random Forest proved to be the best baseline model. Random Forest resulted in the following metrics, AUC = 0.926, 95% CI = [0.910,0.940], accuracy score = 0.968, F1 score = 0.915. It can be observed that the results of the proposed approach are far better than the results of the best baseline model.

### Shapley Value Analysis

Figure 5 illustrates the SHAP value results. Based on this figure, HOSP_LOS had the most significant impact on the prediction of mortality for ICU patients with Sepsis-3, followed by ICU Length of Stay (ICU_LOS) and age. Intriguingly, the relationship between HOSP_LOS and mortality appears to be negative, suggesting that extended hospital stays may correlate with lower mortality rates among ICU patients with Sepsis-3. Furthermore, the analysis indicates a positive contribution of ICU_LOS to mortality prediction, as evidenced by the proliferation of red dots towards the right. This implies that prolonged ICU stays may elevate mortality risks for ICU patients with Sepsis-3. Moreover, age and the Elixhauser Comorbidity Index (ELIXHAUSER_HOSPITAL) exhibit positive associations with prediction accuracy. This aligns with intuition, as advanced age and greater comorbidity burden typically elevate patient vulnerability and subsequent mortality risks. Whereas Minimum of Lactate Level (LACTATE_MIN) and Average Blood Urea Nitrogen level (BUN_MEAN) were the least important features for the prediction of mortality. The relative importance of features as indicated by SHAP values helped us refine our model by focusing on the most impactful variables. For instance, recognizing the significance of HOSP_LOS and ICU_LOS enabled us to ensure these variables were accurately represented and considered in our model tuning process. By leveraging SHAP insights, we could also identify less important features, allowing for potential dimensionality reduction and further optimization of the model’s performance. This approach not only improved the interpretability of our model but also its predictive power by concentrating on the features that matter most.

**Figure 5:**
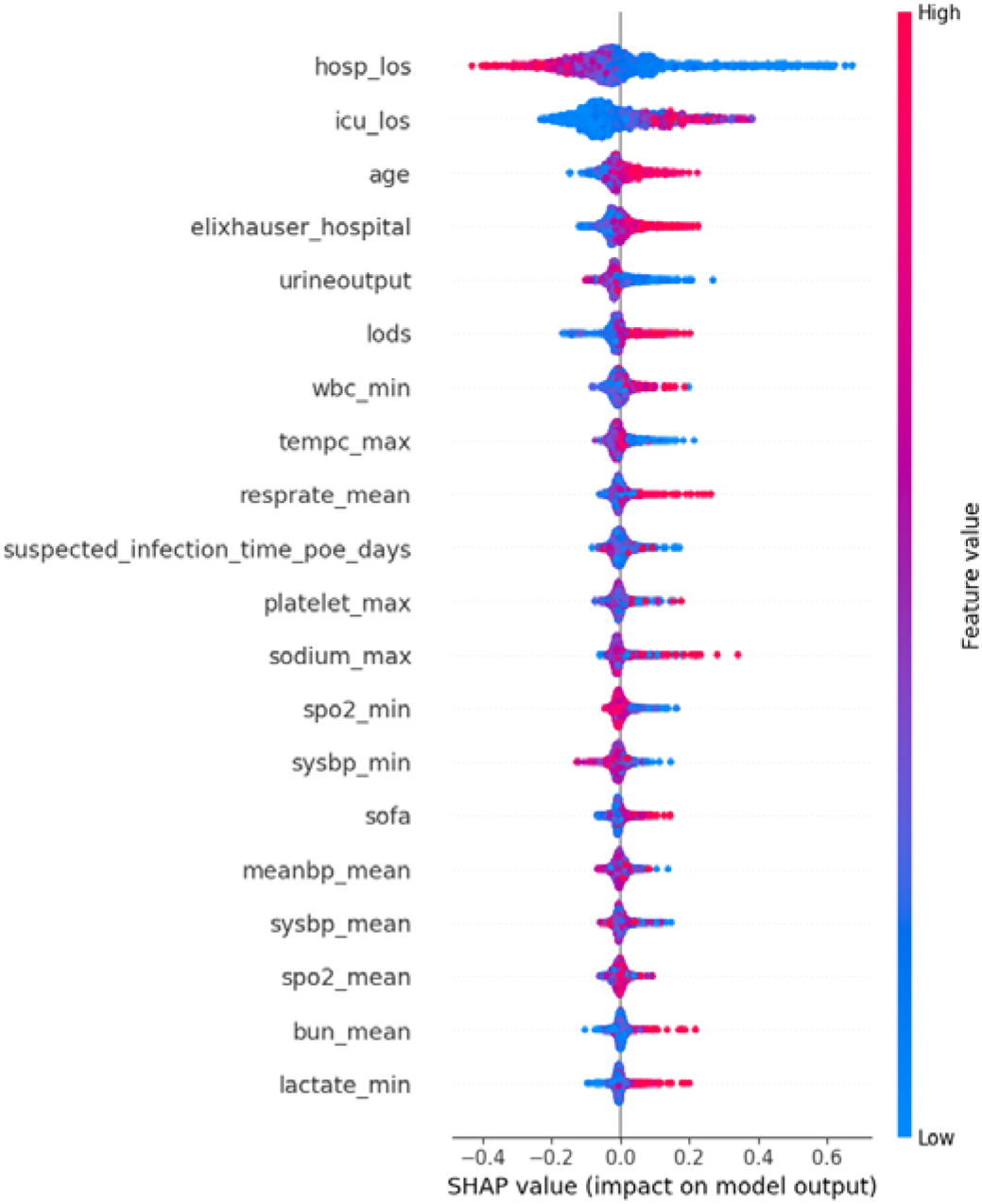
It illustrates the contribution of each feature to the model’s prediction for a specific instance.

This study underscores the critical role of Hospital Length of Stay (HOSP_LOS) in mortality prediction. HOSP_LOS emerged as the most statistically significant factor, with the Mann-Whitney U Test confirming that its absence results in significantly different AUC scores. Unlike ICU Length of Stay (ICU_LOS) and age, which are positively correlated with mortality, HOSP_LOS showed a negative correlation, suggesting that longer hospital stays allow for more effective treatment and management, thus improving survival rates.

Incorporating HOSP_LOS into our model notably enhanced the AUC metric and provided deeper insights into Sepsis-3 patient outcomes. Unlike static indicators like ICU_LOS and age, HOSP_LOS is a dynamic variable that reflects treatment effectiveness, thereby enhancing the model’s accuracy and flexibility. This feature complements other significant variables, making the predictive model more comprehensive and reliable.

### LIME Analysis

In addition to SHAP, we employed Local Interpretable Model-agnostic Explanations (LIME) to provide interpretability at the individual prediction level [34]. LIME explains individual predictions by approximating the black-box model with an interpretable local model around the prediction of interest.

For example, in one of the predictions, four features significantly contributed towards predicting Class 0. These features included age, urineoutput, lods, and resprate_mean, with specific values of 0.30, 0.04, 5.00, and 0.35 respectively. Each of these features influenced the prediction towards one of the classes. (see Figure 6).

**Figure 6:**
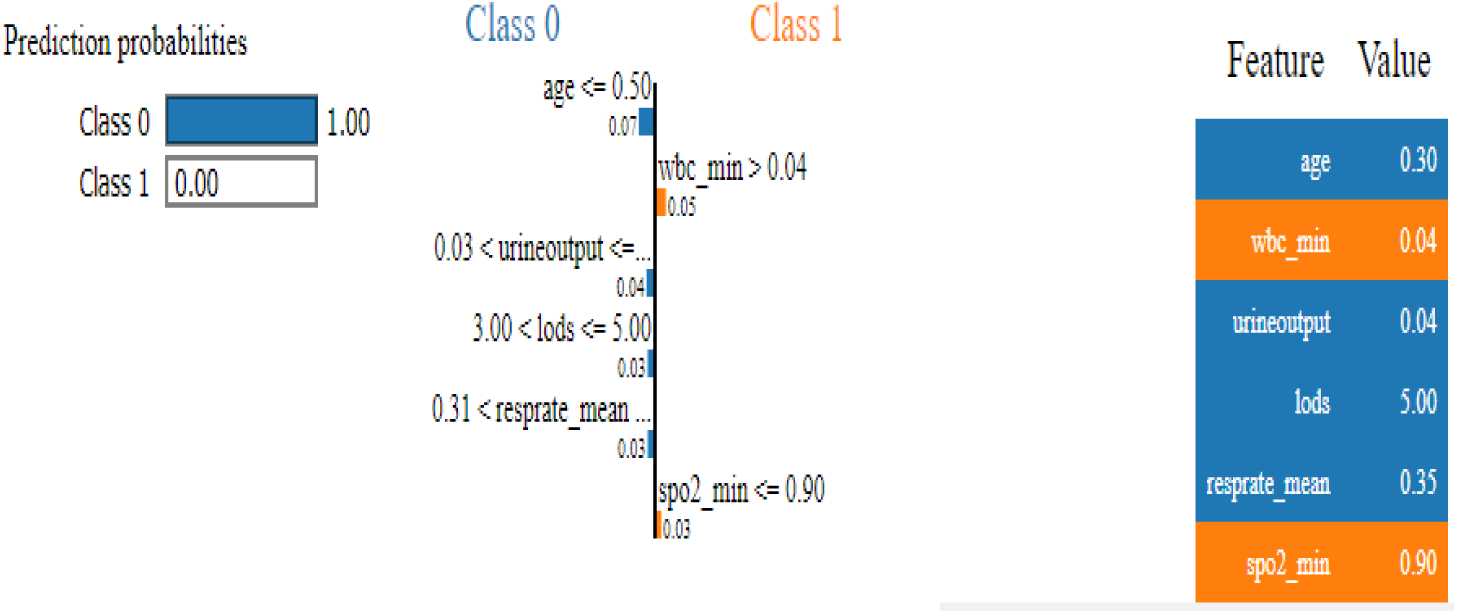
LIME explanation of feature contributions for an individual prediction.

Additionally, spo2_min at 0.90 and wbc_min at 0.04, exactly on the threshold, contributed towards Class 1. While age being 0.30, which is less than 0.50, contributed towards Class 0; wbc_min at 0.04, above the threshold, contributed towards Class 0; urineoutput within the range of 0.03 to 0.04 further contributed towards Class 0; lods at the upper boundary of the 3.00 to 5.00 range added to the likelihood of Class 0; and resprate_mean at 0.35, which is above 0.31, provided additional support towards Class 0. Aniongap_min at 0.18, below the threshold of 0.21, also contributed towards Class 0. These feature values collectively led to a strong prediction of Class 0, illustrating their significant roles in the model’s decision-making process for this instance.

Using LIME, we could explain and visualize how specific features influenced individual predictions, adding another layer of interpretability to our model. The combination of SHAP and LIME provides a comprehensive understanding of both global feature importance and local prediction dynamics, enhancing the overall transparency and trustworthiness of the model.

### LightGBM’s Learning Curve Analysis

Additionally, to evaluate LightBGM’s ability to handle unseen data, its learning curve has been plotted by software Python. Binary error, the proportion of incorrectly classified instances in the dataset, is decreasing as boosting rounds, sequential training of individual decision trees, rises, which is implying model’s classification accuracy. The convergence of both the training and testing curves has been observed. As depicted in Figure 7, a decreasing training error accompanied by a decreasing testing error indicates that LightGBM is learning effectively without overfitting.

**Figure 7:**
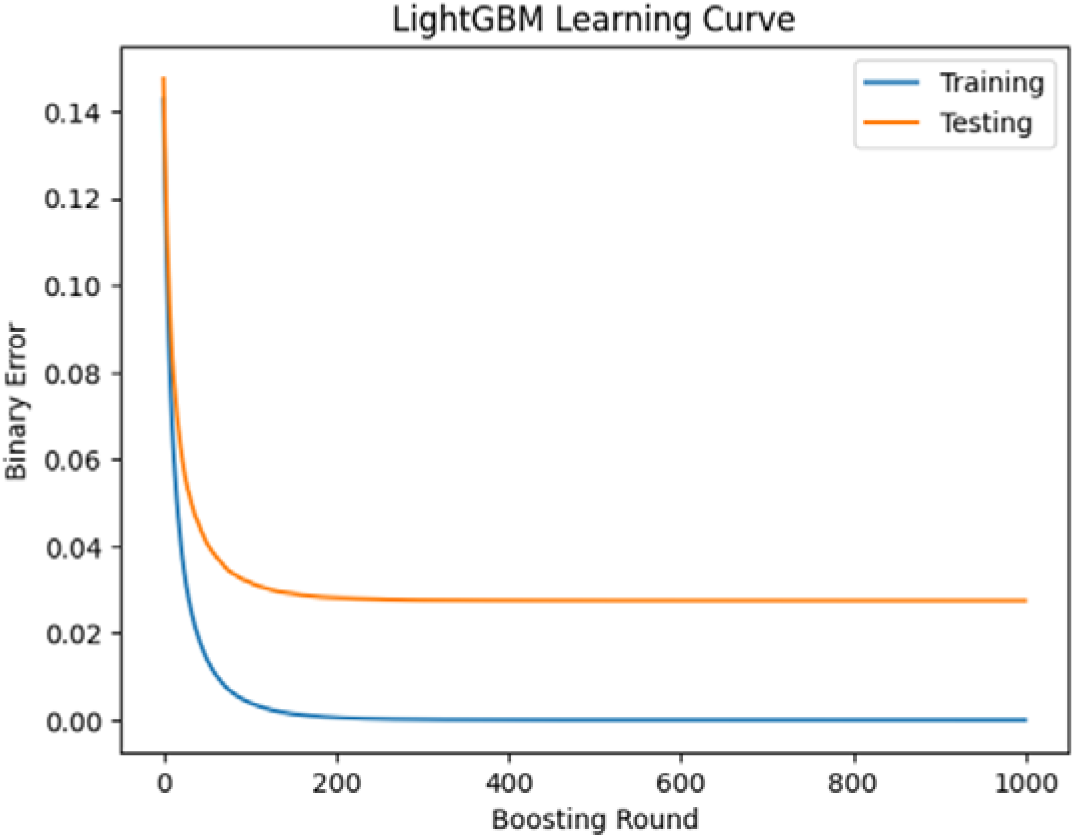
Error curve for training and validation data as tested on the LightGBM model.

## DISCUSSION

### Existing Model Compilation Summary

Several methods have been developed to predict mortality for ICU patients with Sepsis [16,19]. However, only one study focuses on Sepsis-3 using the MIMIC-III database [18], proposing XGBoost with an AUC of 0.857 (95% CI [0.839-0.876]).

In our study, novel data preprocessing techniques have been utilized. Bootstrapping, a statistical resampling method, is applied to augment the limited sample size, doubling our dataset’s size to 9118 samples.

Our novel feature selection approach, Entropy Analysis using Decision-Tree, has allowed our proposed model to produce more accurate and robust results. Our approach has resulted in an almost 15% Improvements of AUC metric. Our proposed model, LightGBM, has distinct features such as Gradient-based One-Side Sampling (GOSS) and Exclusive Feature Bundling (EFB), enabling it to be more computationally efficient.

In addition, our approach has also enabled us to find the most statistically significant factor in prediction of mortality, which is HOSP_LOS. Mann-Whitney U Test is also performed to prove that the absence of HOSP_LOS results in statistically different AUC score, and this feature is not included in the best existing literature [18].

Although the existing proposed methodology in the literature was successful in predicting 30-day mortality for ICU patients of Sepsis-3, it possessed several drawbacks. First, they have failed to consider more efficient feature engineering techniques such as Entropy Analysis, thus it ignored the most significant feature in predicting mortality which is HOSP_LOS. Secondly, the existing literature has a wide confidence internal for its AUC metric. A wide confidence interval can be a consequence of an inadequate sample size used for evaluation. To provide a clearer comparison, we have summarized the differences in methodologies and results between our study and the best existing literature in Table 4.

**Table 4:** Comparative Analysis Between Our Study and the Best Existing Study.

|  | Hou et al. (2020) | Our Study |
| --- | --- | --- |
| Proposed Model | XGBoost | LightGBM |
| AUC | 0.857 | 0.983 |
| 95% CI for AUC | 0.839-0.876 | 0.980-0.990 |
| Feature Engineering Techniques | Stepwise Analysis | Entropy Analysis using Decision Tree |
| Top 5 Significant Features | Urine output, Lactate min, Bun mean, sysbp min, inr max | hosp_los, icu_los, Age, Elixhauser hospital, Urine output |

Our proposed approach offers several advantages over prior research: (a) Novel preprocessing and feature engineering techniques significantly enhanced model performance, achieving substantial improvements over existing literature. (b) Identification of a crucial feature, HOSP_LOS, which was overlooked by other studies but is statistically significant in predicting mortality. (c) Utilization of LightGBM and Grid Search CV, which are more effective for handling large-scale, high-dimensional datasets with better computational efficiency. These advancements resulted in nearly a 10% improvement over existing models, highlighting the efficacy of our methods in enhancing predictive performance and advancing mortality prediction in the medical domain.

### Practical Applications

Our research has several practical applications. Accurate prediction of 30-day mortality for Sepsis-3 patients allows hospitals to better allocate ICU resources, directing critical care to those at higher risk. This can lead to more personalized and timely interventions, improving patient outcomes and reducing mortality rates. The inclusion of features like Hospital Length of Stay (HOSP_LOS) enhances the model’s predictive power, aiding clinicians in making informed decisions about patient care. Additionally, the streamlined preprocessing methodology reduces the number of relevant features, making the model more efficient and less resource intensive. Furthermore, our methods can be adapted to predict outcomes for other critical conditions in ICU settings, broadening the research’s impact across various medical scenarios.

### Study Limitation

Our study has several main limitations. Firstly, the lack of external datasets like MIMIC-III restricts our ability to validate the model’s effectiveness. Future research should aim for external validation with comprehensive and contemporary data from multiple institutions, ensuring robustness across diverse healthcare environments. The MIMIC-III database is over 10 years old and lacks historical information, introducing potential biases. Utilizing newer databases reflecting current clinical practices and technologies will enhance model accuracy and applicability.

Despite using robust entropy-based feature selection, relevant features might still be missed. Continuous collaboration with clinical experts is essential to ensure comprehensive feature consideration. Additionally, the MIMIC-III database originates from a single medical center, which may introduce biases due to variations in patient demographics and treatment protocols across different institutions.

Model overfitting remains a concern, as high performance on training data may not fully translate to real-world settings. Extensive external validation and potential model retraining are necessary to confirm generalizability. Furthermore, the temporal changes in clinical practices, treatment protocols, and healthcare technology, given the MIMIC-III dataset spans from 2001 to 2012, could affect the model’s applicability to current patient populations.

Data imputation, while necessary, can introduce biases. Future studies should explore alternative imputation methods and their impacts to ensure robustness. Although bootstrapping increased the sample size, the final cohort of 9118 patients may still be relatively small for certain sub-analyses. Larger datasets could improve the model’s accuracy and reliability.

Addressing these threats in future research will enhance the robustness and applicability of our predictive model for ICU patients with Sepsis-3. While the current study advances the prediction of 30-day mortality for Sepsis-3 patients, acknowledging and addressing these limitations is crucial. Future research should focus on external validation and incorporating more recent and diverse datasets to improve the generalizability and reliability of the predictive model.

## CONCLUSION

Utilizing bootstrapping, Entropy Analysis via Decision-Tree algorithms, data imputation strategies, and model optimization techniques not only enhances predictive model accuracy and robustness but also improves generalization to unseen data. This is evident from notable improvements in AUC and Accuracy metrics, outperforming existing methodologies. These advancements underscore the importance of feature selection for computational efficiency and highlight the potential of ML as a valuable tool in the dynamic ICU environment, where precise predictive models are crucial.

In future studies, the proposed approach could be validated using datasets from other healthcare systems to ensure its applicability and robustness across different populations and settings. Moreover, incorporating the comprehensive information available in the MIMIC-III dataset, such as clinical notes and images, as inputs for models holds promise for further research and exploration. These rich data sources could provide additional context and features, potentially leading to even more accurate and reliable predictive models. By leveraging diverse data types and sources, we aim to develop models that not only perform well in controlled settings but also demonstrate strong generalizability and reliability in real-world clinical environments.

Ultimately, through ongoing research efforts and the integration of varied datasets and advanced data types, we anticipate further enhancing the model’s generalizability and performance. This continuous improvement is essential to developing reliable tools that can aid clinicians in making informed decisions, thereby improving patient outcomes in intensive care units and beyond.

## Data Availability

All data produced are available online at https://physionet.org/content/mimiciii/1.4/

## ABBREVIATIONS

ML: Machine Learning
ICU: Intensive Care Unit
SAPS-II: Simplified Acute Physiology Score-II
SOFA: Sequential Organ Failure Assessment
AUROC: Average Area Under the Receiver Operating Characteristic Curve
LIME: Local Interpretable Model-agnostic Explanations

## Acknowledgments

The authors extend their gratitude to the developers of MIMIC-III for offering a detailed and comprehensive public EHR dataset.

## Author information

### Authors and Affiliations

Department of Industrial and Systems Engineering, University of Southern California (USC), Andrus Gerontology Center, 3715 McClintock Ave GER 240, Los Angeles, CA 90089

Yu Z, Ashrafi N, Li H, Pishgar M

Department of Health Science, California State University, Long Beach (CSULB), 1250 Bellflower Blvd, Long Beach, CA 90840, United States of America

Alaei K

## Authors’ contributions

Z.Y., M.P.: Involved in all aspects of this study. N.A., H.L.: Major revision of the manuscript. Expert insights were provided by K.A.

## Declaration

### Availability of data and materials

The MIMIC-III database which was used during the current study is publicly available. The Medical Information Mart for Intensive Care III (MIMIC-III) is a comprehensive dataset, available to the public via https://physionet.org/content/mimiciii/1.4/

### Ethics approval and consent to participate

The dataset used to support the conclusions of this article is sourced from the Medical Information Mart for Intensive Care version III (MIMIC-III). As this database is public and de-identified, informed consent and Institutional Review Board approval were not required. All procedures followed the relevant guidelines and regulations.

### Consent for publication

Not applicable.

